# Autoimmune Encephalitis Presenting with Malignant Catatonia in a 40-Year-Old Male Patient with Covid-19

**DOI:** 10.1101/2020.07.23.20160770

**Authors:** Jan Mulder, Amalia Feresiadou, David Fällmar, Robert Frithiof, Johan Virhammar, Annica Rasmusson, Elham Rostami, Eva Kumlien, Janet L. Cunningham

## Abstract

Acute malignant catatonia with autonomic instability developed in a previously healthy man with PCR-verified SARS-CoV-2. CT and MRI were normal, EEG showed slowing and cerebrospinal fluid showed a subtle indication of inflammation. There were no signs of pathology in other organs. ^18^F-FDG-PET conveyed high bilateral uptake in the striatum. While commercial tests were negative, immunohistochemical staining of mouse brain revealed anti-neuronal IgG antibodies against neuronal targets in the hippocampus, thalamus, striatum and cortex. Early treatment with plasmapheresis and corticosteroid reversed disease progression and may have prevented large-scale neurological damage. We are not aware of other types of encephalitis with such distinct pyramidal tract symptoms and raise the possibility that this may be a novel form of autoimmune encephalitis induced by infection with SARS-CoV-2.

## INTRODUCTION

A 40-year-old man showed acute debut of agitation, grimacing, repetitive speech and movements (verbigeration and stereotypies) that started earlier the same day. His Glasgow coma scale score was 14. Because his behaviour was bizarre, disorganised, hyperkinetic and uncooperative, a decision was made for involuntary commitment. He had been experiencing high levels of stress in recent months as his business was struggling. Twenty-two days before he developed Covid-19 related respiratory symptoms, dyspnoea and fatigue, which did not require hospital care and he had tested positive for SARS-CoV-2 RNA in a nasopharyngeal swab (day 14; Figure 1A). Anosmia and ageusia were not present. Before admission, he had suffered from a headache. On admission (day 22), he no longer had respiratory symptoms but did have a fever (38.4 °C). He made no eye contact, reflexes were normal and Babinski’s sign was absent. Brain CT, metabolic parameters and blood tests were unremarkable. The patient was lightly sedated, with midazolam followed with dexmedetomidine. Neuroleptics were not used. Treatment with antibiotics and acyclovir was initiated until the tests excluded bacterial infection and herpes encephalitis.

**Figure 1:**
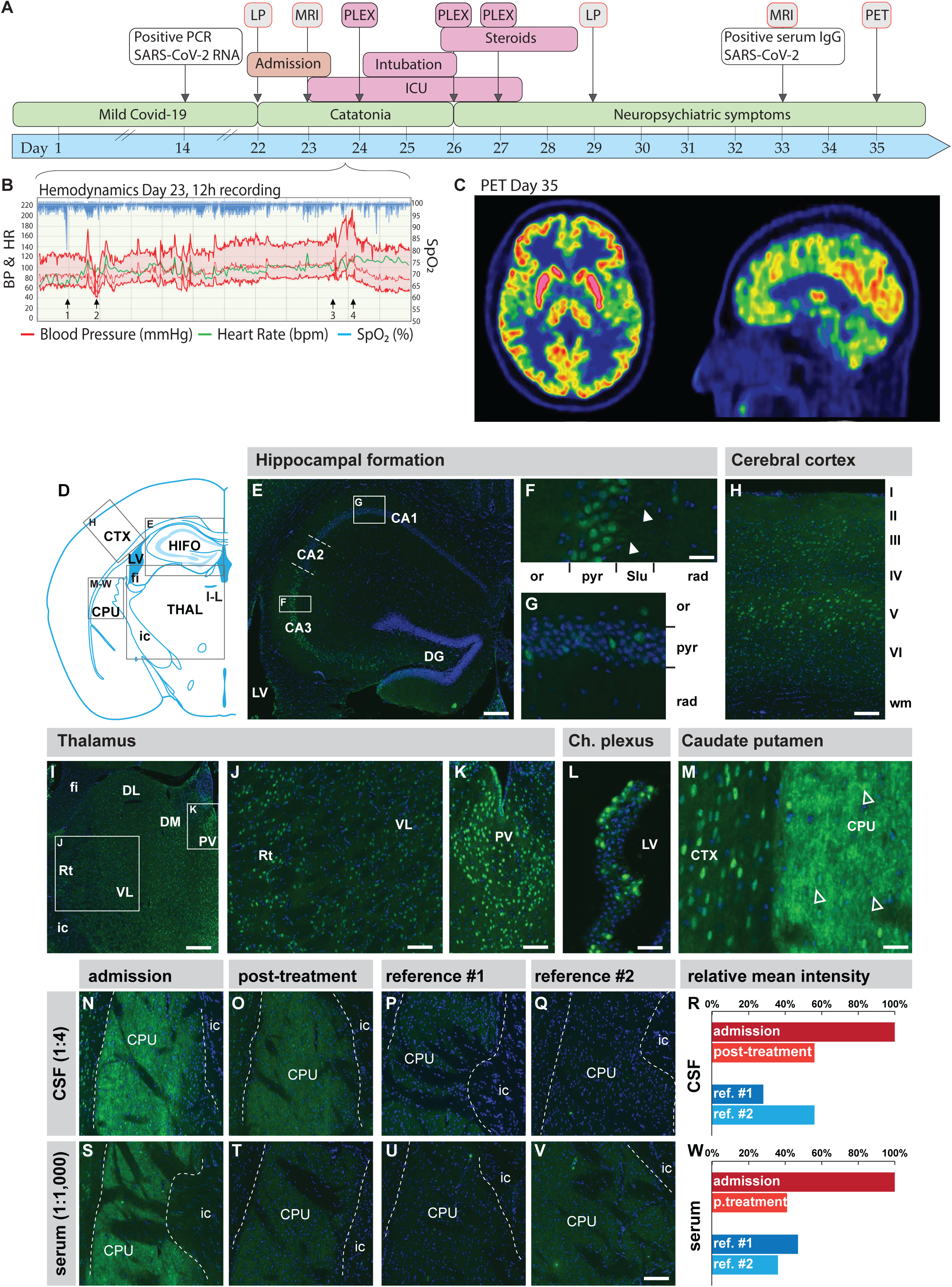
Panel A shows a timeline of evaluations, treatments and progression of symptoms. PLEX, plasmapheresis; ICU, intensive care unit; LP, lumbar puncture; MRI, magnetic resonance imaging, PET-positron emission tomography. Panel B shows an original 12-hour chart recording taken before plasmapheresis (PLEX) (Day 23) with peripheral oxygen saturation (% SpO_2_, blue), arterial blood pressure (BP, red) and heart rate (HR, green). The vertical bars represent hours. The patient was initially sedated with an intravenous dexmedetomidine infusion. A hypoxic event of unknown origin was noted (arrow 1). Blood pressure and heart rate were generally variable with episodes of hypotension associated with hyperlactatemia (arrow 2) and hypertension (arrow 3). Hypotension was treated with intravenous crystalloid fluid and noradrenalin. Induction of anaesthesia and endotracheal intubation (arrow 3) was followed by hypertension (arrow 4) that persisted after treatment with bolus doses of propofol, morphine, clonidine and labetalol. After endotracheal intubation, this treatment regimen was changed to propofol, clonidine and morphine. Panel C shows a brain PET scan 60 minutes after injection of 209 MBq of ^18^F-FDG 13 days after plasmapheresis treatment. The uptake in the caudate nucleus and putamen is bilaterally more prominent than the uptake in the cortex. This observation could represent a global cortical decrease in metabolism or symmetrical hypermetabolism of the striatum. On visual inspection, there were no regions with focal cortical anomalies. Panel D shows profiling of IgG immunoreactivity on mouse brain tissue. CSF (diluted 1:4) collected at the first lumbar puncture revealed a moderate to strong immunoreactivity in neuron-like cells in all investigated brain regions. In the hippocampal formation the strongest immunoreactivity was observed in the CA3 pyramidal neurons (E) labelling the nucleus, soma and proximal dendrites in the stratum lucidum (arrowheads in F). In other regions of the hippocampal formation IgG immunoreactivity was found in a small number of neuron-like cells in the pyramidal layer, the oriens layer (G) and the polymorph layer of the dentate gyrus (E). A similar neuronal staining pattern was observed in the somatosensory cortex (H), with the strongest labelling of pyramidal neurons in layer V and the thalamus (I) and the strongest labelling of neurons in the paraventricular nucleus and reticular nucleus compared to dorsolateral and ventrolateral thalamic nuclei (J and K). A subpopulation of ependymal cells lining the dorsal 3^rd^ ventricle and choroid plexus (L) reveals nuclear and cytoplasm staining. The highest level of immunoreactivity was found in neurons and neuropil in the caudate-putamen (M). Note the dotted spine-like staining pattern (arrows in M). CSF (N) and serum (S) from before treatment compared to after treatment (O and T) revealed less of the same pattern of immunoreactivity with a remarkably reduced intensity when compared. Reference was CSF (diluted 1:4; P and Q) and serum (diluted 1:1,000 U and V) from two age-matched men with Bipolar I disorder in manic phase. Quantification of immunoreactivity before and after treatment is shown for CSF respective serum (R and W). Abbreviations: CA(1-3), cornu ammonis (1-3); CPU, caudate-putamen; CTX, cerebral cortex; ic, internal capsule; DG, dentate gyrus; DL, dorsolateral thalamus; DM, dorsomedial thalamus; fi, fimbria of the hippocampus; HIFO, hippocampal formation; LV, lateral ventricle; or, oriens layer; PV, paraventricular thalamic nucleus; pyr, pyramidal layer; rad, radiatum layer; Rt, reticular nucleus; Slu, stratum lucidum; THAL, thalamus; VL, ventrolateral thalamus.

Lumbar puncture showed high red blood cells, 19000 cells ×10^6^/L, secondary to traumatic lumbar puncture. Cerebral spinal fluid (CSF) cell count indicated pleocytosis with 23 mononuclear and 8 polymorphonuclear cells ×10^6^/L. Signs of blood-brain barrier disruption with elevated albumin levels in CSF of 838 mg/L (reference <400 mg/L) and the CSF/serum quotient was 15.6 (>6.8). Interleukin 6 (IL6) in CSF was high: 102.1 pg/mL (<5 pg/mL). IL6 in plasma, taken a day later, was 30 pg/ml (<7.0). Neurofilament (NFL), glial fibrillary acidic protein (GFAP) and tau protein in CSF were normal. PCR tests were negative in CSF for SARS-CoV-2. Antibodies against anti-neuronal antibodies were negative using a commercial assay (Euroimmun, Germany). Brain MRI was normal.

A few hours later, the patient’s state deteriorated, temperature rose to 39°C and he became mutistic, showing signs of autonomic instability with recurrent episodes of fluctuating heart rate and arterial blood pressure. He also exhibited periods of oxygen desaturation (Figure 1B). When hypertension ensued, it was difficult to treat, despite high doses of clonidine and labetalol. Plasma lactate varied between 0.6 and 8 mmol/L but myoglobulin but CKMB remained normal. His pupil size, reaction to light and the oculo-cephalic reflex was normal. Slow, horizontal roving eye moments were noted. He displayed decorticate posturing, increased tonus, resisted movement of arms and jaw but had normal tonus in the legs. Hyperreflexia was present with bilateral foot clonus and Babinski’s sign but no neck stiffness. Anaesthesia was induced to facilitate endotracheal intubation. The patient was thereafter continuously sedated with propofol and clonidine. D-Dimer (1.2mg/L) was slightly elevated but there were no signs of thromboembolic events. Respiration and cardiovascular function remained stable. Continuous EEG monitoring showed non-specific slowing with left hemisphere predominance without epileptiform activity. An episode of asystole with spontaneous recovery, episodes of bradycardia of 27 bpm and repeated p-waves without QRS waves were interpreted as atrioventricular block III and the patient was monitored for 3 days with telemetry.

## RESULTS

Although the diagnosis remained uncertain, post-infectious autoimmune encephalitis was suspected. Plasmapheresis was therefore initiated and repeated three times over 4 days. On day 26 and after two courses of plasmapheresis, the patient was extubated and was autonomically stable. Eye movement was normalised, hyperreflexia was less prominent and bilateral Babinski’s sign persisted. This case shared features with a recently published steroid-responsive case^1^ and treatment was therefore initiated with 1g methylprednisolone per day and discontinued after 3 days because of insomnia and elevated mood.

On day 28, the patient showed a dramatic improvement. He was awake, oriented, communicative but had no memories from the past days. He had visual hallucinations and cognitive symptoms that improved over the coming weeks. The patient’s EEG was normal.

A second lumbar puncture, performed on day 29, showed pleocytosis, 10 mononuclear cells and 1 polymorphonuclear cell ×10^6/^L, elevated IgG levels and IgG index, as well as two oligoclonal bands in CSF not represented in serum, indicating intrathecal production of antibodies. PCR tests were still negative in CSF for SARS-CoV-2. IL6 in CSF was normalised at 4.8 pg/ml. GFAP and Tau remained normal but NFL was increased to 1030 ng/L (<890). A second MRI and a standard neurological exam on day 31 were normal, and no abnormal eye movements. The hallucinations were less frequent. He described increased emotional lability and mental fatigue with disturbed short-term memory and decision making. He also found it challenging to recognise the voices and faces of acquaintances. Serology taken on day 33 was strongly positive for IgG against SARS-CoV-2. ^18^F-FDG PET scan, day 35 (13 days after treatment initiation), showed high bilateral uptake in the striatum (caudate nucleus and putamen) when compared to the cortex (Figure 1C).

## EVIDENCE OF AUTOIMMUNITY

Using immunohistochemistry we detected IgG autoantibodies against mouse brain neuronal proteins in serum and CSF at admission (Figure 1D-W). Immunoreactivity was observed in all brain regions investigated (cerebral cortex, hippocampal formation, thalamus, caudate-putamen) with strongest signal observed in the nuclear and perinuclear compartments without reactivity in the nucleoli. Neuronal labelling intensity was strongest in the CA3 in the hippocampal formation, layer V in the somatosensory cortex and the paraventricular and reticular nucleus in the thalamus. A subset of ependymal cells located in the ventricle wall and choroid plexus revealed strong immunoreactivity of the (peri)nuclear compartment and cytoplasm. Immunoreactivity of neuropil was most intense in the caudate-putamen, revealing neuronal processes and spine-like structures. Post-treatment IgG immunoreactivity in the (peri)nuclear compartment and neuropil was notably reduced, reaching the levels of reference CSF and serum.

## DISCUSSION

This report is a case of suspected autoimmune encephalitis in a patient with Covid-19 who presented with excited catatonia progressing to malignant catatonia with autonomic instability Signs of autoimmune encephalitis were present but this case did not initially meet the proposed criteria.^2^ Standard radiological findings were normal and the discrete pleocytosis and elevated protein in CSF may have easily been ignored as non-specific. Cases are reported with rapid onset of encephalitis with diffuse corticospinal tract signs in conjunction with Covid-19.^3,4^ Neurological symptoms may also occur as an early Covid-19 manifestation in patients who later develop typical symptoms.^4^ Neurological symptoms in Covid-19 patients include one case of catatonia^5^ as well as mutism.^1^ Autonomic dysfunction after Covid-19 has been reported in a patient with psoriasis.^6^ Several forms of autoimmunity after Covid-19 are emerging and this coronavirus may join other neurotropic viruses as a risk factor for autoimmune encephalitis.^7^

Malignant catatonia is a more severe form of the catatonia spectrum but perhaps less recognised than the typical form with immobility, waxy flexibility and stupor.^8,9^ A recent review identified 124 cases of catatonia in conjunction with infections, of which 38% were viral.^10^. Catatonia may occur in patients with no psychiatric history and is co-occurring with several autoimmune conditions, especially NMDAr encephalitis.^10^ The staining, clinical presentation and radiology are consistent with a synaptic target highly present in the striatum, where binding leads to increased excitation. Possible mechanisms include disinhibition or activation of excitatory receptors as well as disturbance of excitatory neurotransmitter uptake/clearance or interference with ion homeostasis.

In patients with acute respiratory distress syndrome related to Covid-19, agitation diffuse corticospinal tract signs along with enhanced reflexes and ankle clonus are common findings seen in 67% of cases^3^. In the same study, dysexecutive symptoms were observed in fewer cases (37%).^3^ The suggested causes are critical illness-related encephalopathy, elevated cytokines, or the effect of withdrawal of medication. Autoimmunity in these cases, though speculated, has not yet been reported.

The present case was not classed as a critical illness and IL6 was only moderately elevated. Normal MRI and NFL levels in CSF did not support neuronal injury. The^18^F-FDG PET findings in the striatum are consistent with increased metabolism in local neurons, glial cells, or both. Striatal hypermetabolism has been previously reported for autoimmune encephalitis caused by antibodies against NMDAr and VGKC encephalitis.^11-13^

The swift introduction of treatment with plasmapheresis and high dose steroids may have prevented the development of more severe neurological damage.. The treatment reduced autoreactive IgG in both serum and CSF but may also have other actions.^14^

We conclude that malignant catatonia with potentially life-threatening autonomic instability can occur in patients with Covid-19 and is likely a form of para-infectious autoimmune encephalitis. This condition responded with dramatic improvement and there was no evidence of structural brain damage. We are not aware of other types of encephalitis with such distinct pyramidal tract symptoms and raise the possibility that this may be a novel form of autoimmune encephalitis induced by infection with SARS-CoV-2.

## Data Availability

Available upon request.

## AUTHOR CONTRIBUTIONS

AF and JLC collected the clinical data and developed the theoretical framework; JM performed the immunohistochemistry experiments; DF analysed the radiological data; EK interpreted the neurophysiology results; EA and AR aided in sample acquirement; JLC designed the study and drafted the manuscript; AR, JM, JV and JLC designed the figure. All authors helped interpret the results and worked on the manuscript. All authors discussed the results and commented on the manuscript.

## ETHICS

This study was approved by the Swedish Ethical Board. The patient has approved the publication of his case and signed an informed consent form. All animal experiments conformed to the European Communities Council Directive (86/609/EEC) and in accordance with Swedish laws and regulations, and all experiments were approved by the local ethical committee (Stockholms Norra Djurförsöksetiska Nämnd N183/14).

